# Synthetic apparent diffusion coefficient for high *b*-value diffusion weighted MRI in Prostate

**DOI:** 10.1101/19008201

**Authors:** Prativa Sahoo, Russell Rockne, Jung Alexander, Pradeep K Gupta, Rakesh K Gupta

## Abstract

**Purpose:** It has been reported that diffusion weighted imaging (DWI) with ultrahigh b-value increases the diagnostic power of prostate cancer. DWI imaging with higher b-values is challenging as it commonly suffers from low signal to noise ratio (SNR), distortion and longer scan time. The aim of our study was to develop a technique for quantification of apparent diffusion coefficient (ADC) for higher b-values from lower b-value DW images.

**Materials and Methods:** Fifteen patient (7 malignant, 8 benign) with prostate cancer were included in this study retrospectively with the institutional ethical committee approval. All images were acquired at 3T MR scanner. The ADC values were calculated using mono-exponential model. Synthetic ADC (sADC) for higher b-value were computed using a log-linear model. Contrast ratio (CR) between prostate lesion and normal tissue on synthetic DWI (sDWI) was computed and compared with original DWI and ADC images.

**Results:** No significant difference was observed between actual ADC and sADC for b-2000 in all prostate lesions. However; CR increased significantly (p=0.002, paired t-test) in sDWI as compared to DWI. Malignant lesions showed significantly lower sADC as compared to benign lesion (p=0.0116, independent t-test). Mean (±standard deviation) of sADC of malignant lesions was 0.601±0.06 and for benign lesions was 0.92 ± 0.09 (10^−3^mm^2^/s).

**Discussion / Conclusion:** Our initial investigation suggests that the ADC values corresponding to higher b-value can be computed using log-linear relationship derived from lower b-values (b≤1000). Our method might help clinician to decide the optimal b-value for prostate lesion identification.

## Introduction

In the past few years, the use of diffusion weighted magnetic resonance imaging (DWI, MRI) for disease detection and characterization has increased substantially. For instance, several studies have assessed the importance of DWI derived apparent diffusion coefficient (ADC) in characterization of prostate cancer aggressiveness(1–4). Quantification of ADC is based on at least two diffusion weighted (DW) images with different b-values. In general, a mono-exponential fit between the natural logarithm of the signal intensity against the b-value yields the ADC. In literature, various other mathematical models have been suggested for ADC quantification, such as stretched-exponential, Gaussian and Kurtosis(5,6). However, in prostate, mono-exponential fit for ADC calculation is sufficient to discriminate prostate cancer from normal tissue(5). Moreover, different ADC values can be found in literature due to the variation in the b-value used to compute the ADC (7). Deciding the optimal *b*-value for prostate cancer characterization is an active area of research (8–11). In most DWI studies, b-values of 1000 sec/mm^2^ or less used for prostate cancer detection or evaluation [4,6,7]. Normal parenchyma can show higher signal intensity in DWI with *b*-values of 1000 sec/mm^2^ or less, which can make it difficult to distinguish normal tissue from cancer tissue. It has been reported use of higher *b*-values improves disease visualization and detection by increasing contrast between cancerous and noncancerous lesions(10,12,13). Although the use of higher b-values (>1000 sec/mm2) is desirable, obtaining higher b-value DW images is challenging as it leads to decreased signal to noise ratio (SNR), increased distortion, susceptibility artifact and increased scan time. Computed DWI techniques have been proposed to overcome these difficulties (14–18).

Computed DWI is a mathematical technique, which generates images of higher *b*-values by using at least two different lower *b*-value (*b* ≤ 1000) images. It involves computing the ADC map from two lower b-value DW images by using the following equation:

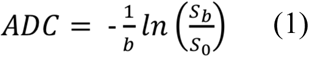

where *S*_0_ is the signal intensity at *b*=0 s/mm^2^. Once ADC for lower *b*-value is known, computed DW images of higher *b*-value can be extrapolated by solving Equation 1 for S_b_,

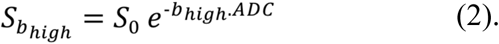

The underlying assumption of computed DWI method is that the ADC is independent of b-values, which contradicts the observation that ADC can vary significantly with the b-value as reported in literature(19,20). Using this technique DW images for higher b-values can be generated but the ADC value for higher b-value cannot be obtained. Computed DWI technique might be useful for the visualization purpose however for quantitative DW image analysis, it might not be sufficient. Therefore, there is a need of methods for generating synthetic ADC maps for higher *b*-values. To the best of our knowledge, methods for creating synthetic ADC maps have not been reported.

The primary objective of this study was to explore the relationship between ADC and b-values and use that relationship to extrapolate synthetic ADC corresponding to higher b-values. A secondary objective was to investigate the feasibility of this technique to improve visualization of lesions in prostate cancer cases for which higher *b*-value DWI may be desirable.

### Theory

Diffusion of water through biological tissue is often quantified using the apparent diffusion coefficient calculated from pairs of b-value DW images using the mono-exponential model (Eq. 1). However, as many studies have demonstrated, the ADC follows a multi-exponential law with respect to higher b-value DWI signal intensity, moreover this multi-exponential behavior is not only related to perfusion artifact (6,7,15,21,22). The multi-exponential behavior depends upon the intra-voxel proton pools that contribute to the signal decay. To overcome the difficulty of making assumptions about the number of intra-voxel proton pools with different diffusion coefficients in biological tissue, Bennett et al. (6) introduced the stretched-exponential model. The stretched-exponential model is described as follows:

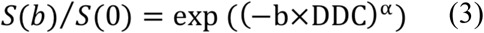

where α represents intra-voxel heterogeneity and DDC is the distributed diffusion coefficient representing the mean intra-voxel diffusion rate, where α=1 is equivalent to the mono-exponential signal decay. Comparing Eq.1 and Eq.3, the ADC computed from mono-exponential model can be written as a function of b:

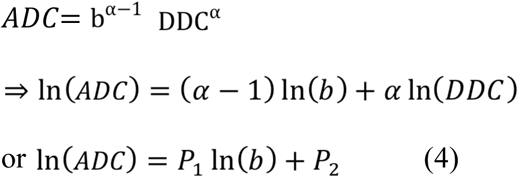

where P_1_ and P_2_ are constants. Therefore, we hypothesized a log-linear relationship between ADC derived from the mono-exponential model and *b*-value. The purpose of this study was to derive the log-linear relation for lower b-value ADCs and use that relationship to extrapolate ADCs for higher *b*-values.

## Material and Method

A total of 15 patients with median age of 62.5 year suspected to have prostate cancer were included in this retrospective study with the institutional ethical committee approval. All the patients’ data were treatment naïve and from a single center. Stereotactic biopsy was performed after the imaging. The Gleason scores (GS) for the biopsies of the malignant tissue were recorded (23). Out of 15 cases, only two patients had GS 7 and five patients had GS 6. The remaining 8 patients were reported as benign. Henceforth we have considered GS 6 and 7 as malignant (N=7) and rest as benign (N=8).

All imaging was performed on 3.0T MR scanner (Ingenia Philips Medical System, Best, The Netherlands). T2-weighted turbo spin echo (TSE) images, covering the whole prostate gland were acquired in the axial plane with parameters: TR-4401ms, TE-120ms, Slice thickness 3mm, number of slice-80, acquisition matrix - 504 × 415, FOV-377 × 377 mm^2^. DWI images were acquired in axial plane with seven different *b*-values (0, 200, 400, 700, 1000, 1500 and 2000 s.mm^-2^), TR-3709ms, TE-77.8ms, Slick thickness-3mm, number of slice 23, acquisition matrix- 92 × 92, FOV- 275 × 275 mm^2^. Acquisition time for all 7 b- value DWI sequence was 3 min 26 sec.

ADC values for different b-values were calculated using mono-exponential model (Eq.1) evaluated voxel-wise. Regions of interest (ROI) of size (15-20 mm^2^) were placed on the transitional zone (TZ) and peripheral zone (PZ) of the prostate for each patient. Variations in the mean ADC value within the ROI with respect to the b-values used for the quantification of ADC were analyzed with a one-way ANOVA test. The log-linear model (Eq.4) was fitted voxel-wise to the lower *b*-value ADCs (ADC_0-400_, ADC_0-700_, ADC_0-1000_) to estimate the model parameters P1 and P2. Synthetic ADC (sADC) calculated from Eq. 4 for b-1500 and b-2000 were extrapolated using the model parameters and compared with the true ADC_0-1500_ and ADC_0-2000_. The error in the sADC at *b*-1500 and *b*-2000 relative to the observed ADC was computed as:

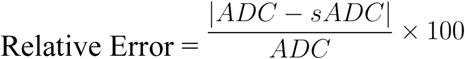

Synthetic DWI (sDWI) images for *b*-1500 and *b*-2000 were generated using DWI of b0 and sADC using mono-exponential model and compared with original DWI_1500_ and DWI_2000_. Contrast ratio (CR) between normal and lesion for DWI and sDWI were computed using CR=(S_cancer_-S_normal tissue_)/(S_cancer_+S_normal tissue_). CR for original DWI and sDWI for *b*-1500 and *b*-2000 sADC values of malignant and benign lesions was assessed by a paired t-test. P values <0.05 were considered as statistical significant. Statistical analysis was performed using Prism (GraphPad Software, Version 7.0).

Regions of interest were placed at normal appearing muscle area and at the lesion on original DWI image and computed DWI image. Two radiologists, one with 10 years of experience and other with more than 20 years of experience blinded to each other and to histological finding, placed the ROIs. For cases with an area suspicious for tumor, ROIs were placed on axial high b-value diffusion weighted images (*b*= 2000 s/mm^2^) on a hyper-intense area suspicious for tumor and a normal intensity area within the gland on the same image. For cases in which the area suspicious for tumor was in the peripheral zone of the gland, the normal intensity region of interest was selected from a location in the peripheral zone on the same image. For cases with no area suspicious for tumor, regions of interest were placed in the relatively hyperintense peripheral zone and in the transition zone -- which is normally hypointense to peripheral zone -- on the same image.

## Results

In one-way ANOVA test, ADC shows highly significant change (p<0.0001) with respect to the b-value, both in transitional zone (TZ) and peripheral zone (PZ) (**Figure 1**) of prostate in all the patient data. This observation supports our initial assumption that the ADC is not constant with respect to *b*-values. The log-linear model gives the best fit to the data (R^2^∼0.9) from the prostate tissue (**Figure 2**).

**Figure 1:**
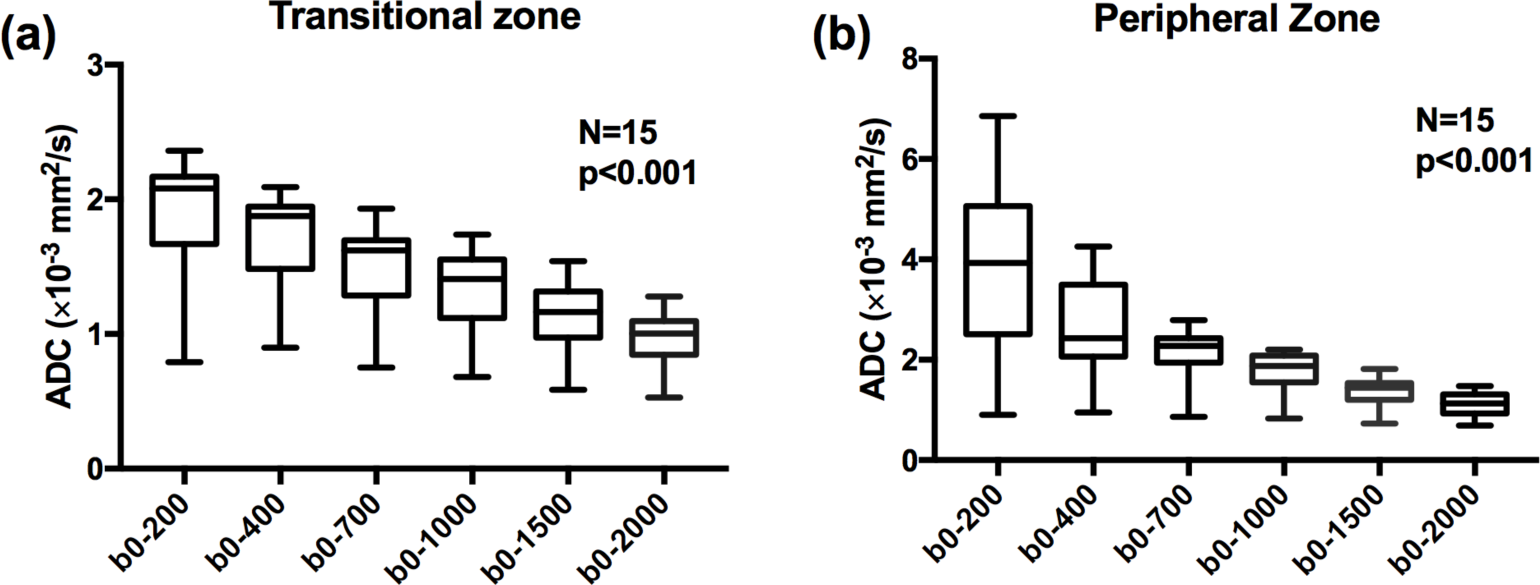
Estimated apparent diffusion coefficient (ADC)using mono-exponential model in the transitional zone (TZ) (a) and peripheral zone (PZ) (b) of prostate. The change in ADC value for each choice of b-value from the other were found to be highly significant with p<0.0001 using one-way ANOVA test in both the regions.

**Figure 2:**
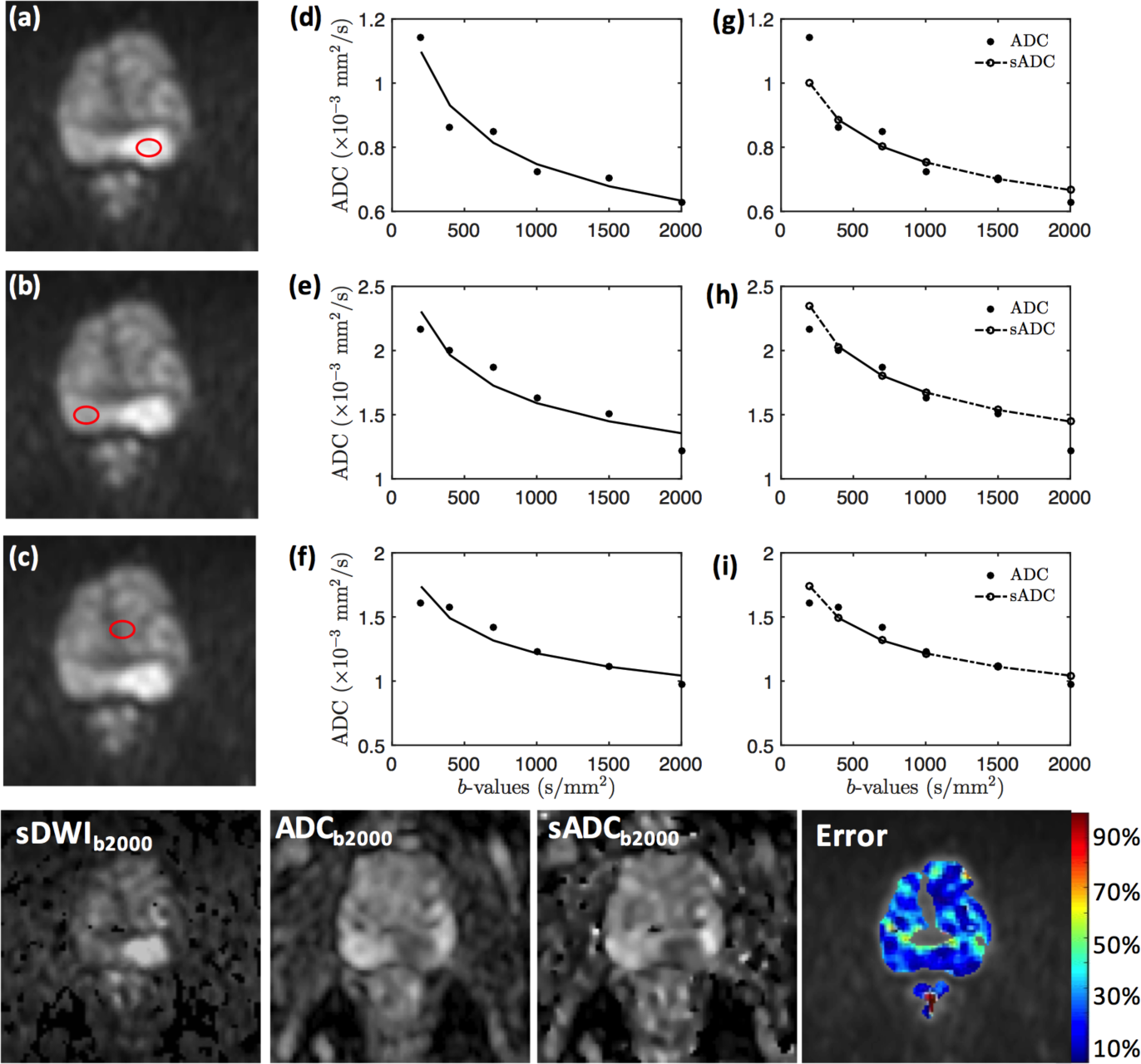
Log-Linear relationship between ADC and b-value. Example of log-linear model fit to targeted tissue of a 69-year-old patient with adenocarcinoma in peripheral zone (PZ) of prostate. Axial DWI images of b-1000 with regions of interest (ROI) in PZ lesion(a), normal PZ (b), normal transition zone (c) and corresponding graphs with b-value (x-axis), ADC (y-axis) and log-linear fit for each ROI (d, e, f). The plots (g, h, i) shows the log-linear model fit to ADC value at b-400, b-700, b-1000(black solid line) and extrapolation of sADC at b-1500 and b-2000(dotted line). Bottom row shows the sDWI, ADC, sADC maps at b-2000 and color coded error map of the corresponding slice.

No significant difference was observed in paired t-test between sADC as compared to actual ADC in the prostate lesions, however the change was significant in the normal tissue (p<0.001) at *b*-2000. Contrast ratio increased significantly between original DWI images and sDWI images (p=0.002) (**Figure 3**).

**Figure 3:**
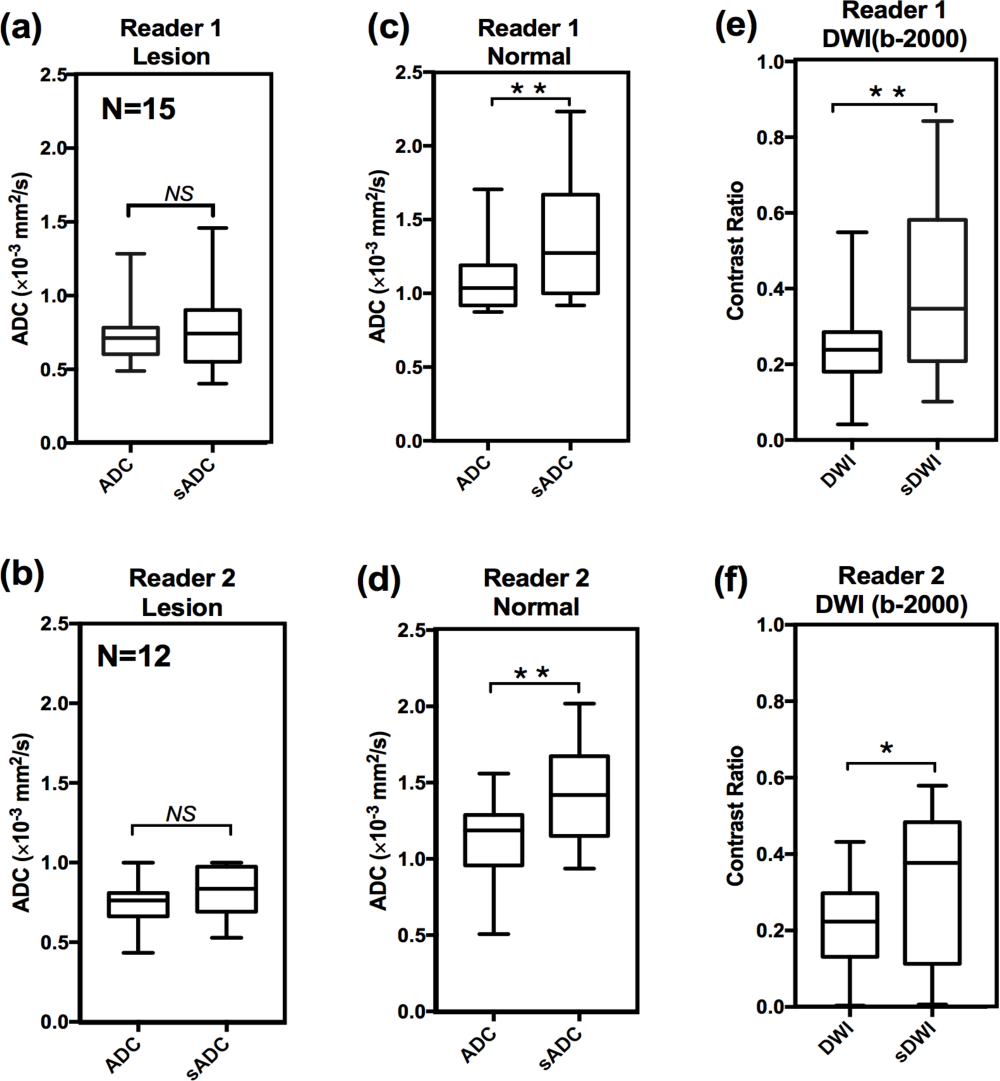
Shows inter-reader variation of ADC and contrast ratio. There was no significant difference between ADC values and synthetic ADC (sADC) values in the lesion (a, b) at b-2000. The difference between ADC and sADC in ROIs placed in normal tissue were significantly different (c, d). However, the contrast ratio of lesion and surrounding normal tissue increased significantly in between DWI and sDWI for b-2000 (e f.). ^⋆^p<0.05, ^⋆⋆^p<0.01 Top row shows the result of Reader 1 (N=15) and bottom row shows that of the Reader 2 (N=12).

Mean sADC of prostate lesions was significantly lower than that of surrounding normal tissue (p<0.001) for *b*-2000 when considered for all data (N=15). A significantly lower sADC was observed using independent t-test in malignant lesions (GS 6,7) as compared to benign lesions (GS<6) (**Figure 4**). In addition, sADC at *b*-1000, *b*-1500 and *b*-2000 was found to be significantly distinguish lesions with GS < 6 from the lesions with GS *≥* 6. The mean sADC value, confidence interval (CI) and the p values are given in **Table 1**

**Table 1:**
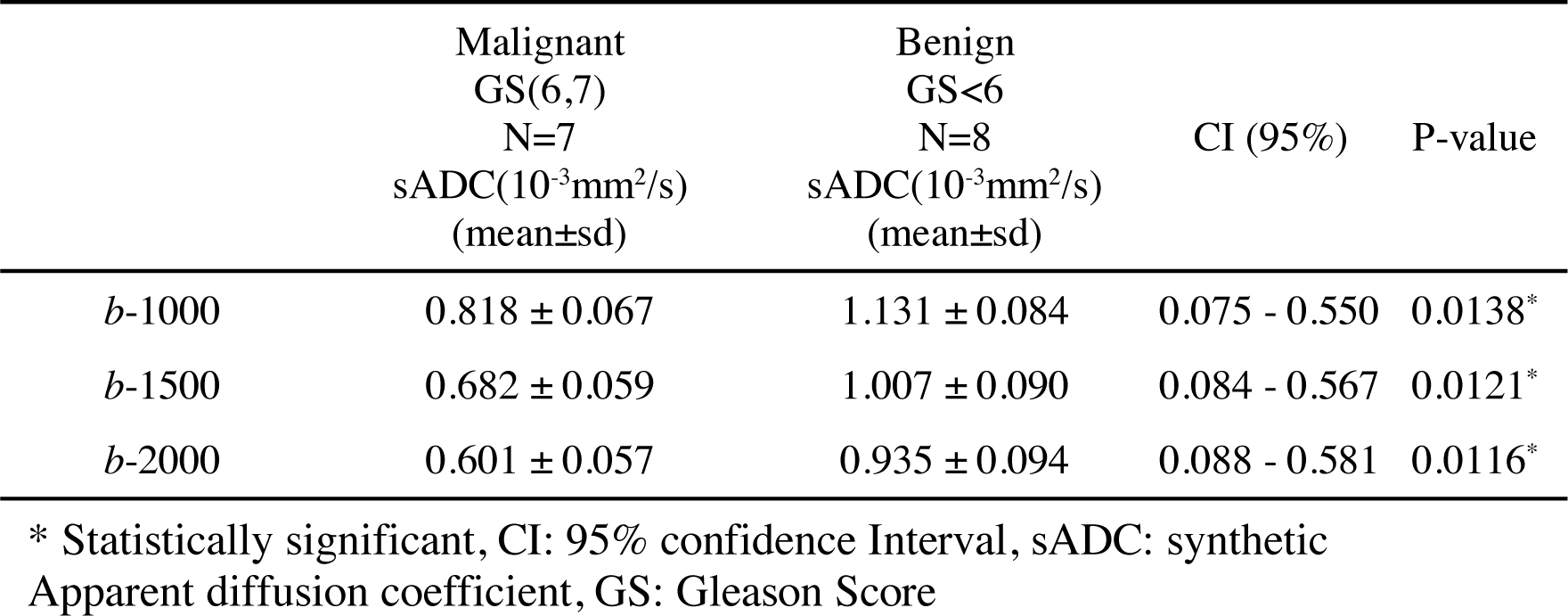
Comparison between sADC values in lesions with Gleason Score (GS)<6 and GS ≥ 6 at b-1000, b-1500 and b-2000.

**Figure 4:**
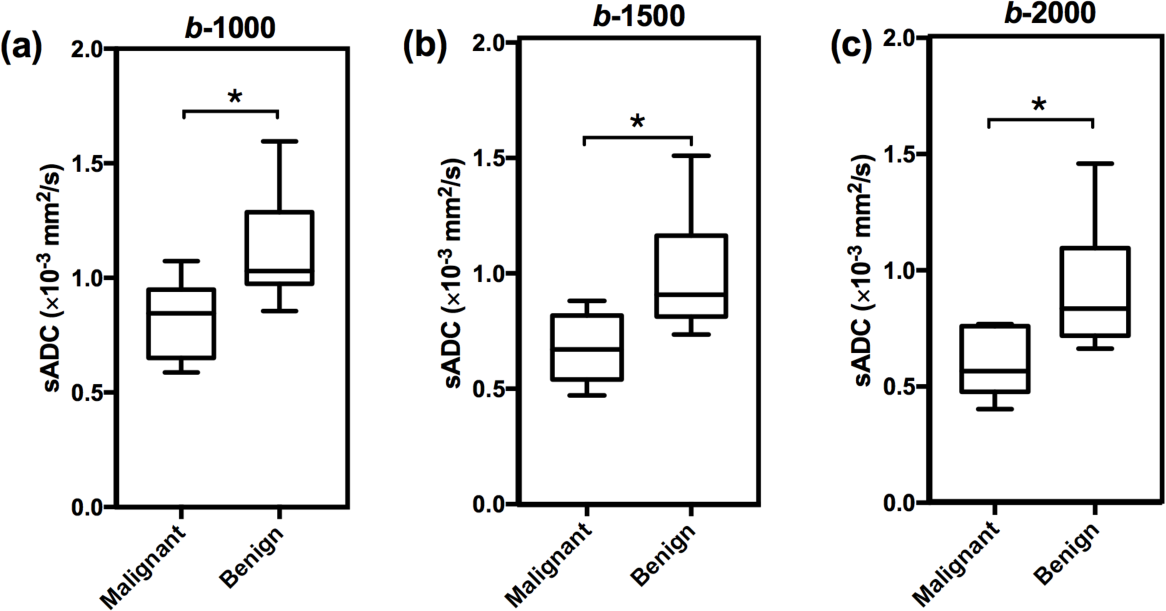
Shows comparison between synthetic ADC (sADC) values of Malignant and benign tissue. Distribution of sADC values for malignant (Gleason score 6,7; N=7) and benign lesions (Gleason score <6; N=8) patients at b-1000 (a), b1500 (b) and b-2000 (c). The center horizontal line indicates the median value. ^⋆^p<0.05

## Discussion and Conclusion

Choice of *b*-values can significantly influence ADC estimation using the mono-exponential diffusion model in prostate, in agreement with variations in ADC found in literature (7,19,20). Our study shows a log-linear relationship between ADC and *b*-values. Using the log-linear relationship derived from ADCs of lower *b*-value (*b*=400, 700 and 1000), ADCs for higher *b*-values (*b*=1500 and 2000) can be extrapolated with a small relative error (10±5)%. Contrast ratio of lesion and normal tissue significantly increases in synthetic DWI images.

The technique of generating synthetic ADC gives clinicians extra degrees of freedom with the choice of b-values. The optimal *b*-value for disease detection depends upon image contrast that is likely to change with tissue type and histological findings. Rather than deciding the optimal *b*-value prior to imaging to get optimal contrast between normal and cancer tissue, the use of synthetic ADC may be able to modify *b*-value and get the optimal image contrast even after imaging. Furthermore, the technique allows extrapolation of ADC values for higher b-values which cannot be obtained by the computed DWI method. However, this technique may not reduce the overall scan time as in our scanning protocol the scanning time to get three different *b*-value (b-400,700 and 1000) is 1 min 39sec and scanning time for one high *b*-value (*b*-2000) is 1 min 5 sec. However, this technique provides a method to obtain DW images and ADC values for a wide range of b-values.

According to the diffusion equation b-value has a [time]^3^dependency, thus a very high b-value can be achieved in clinical scanner with a moderate increase in the echo time (TE). However, the signal loss due to diffusion is a limiting factor at high b-value. The initial signal to noise ratio (SNR) and the tissue diffusion determines how quickly the signal goes below the noise level. As the tissue diffusivity is higher in normal tissue as compared to cancer tissue, normal region signal decay reaches to the noise level at a relatively faster rate. Hence the observed signal at high *b*-values is dominated by the noise and appears to decay at a slower rate. This explains the reason of significant difference in between ADC and sADC values at normal regions. As DWI signal attenuation is exponentially dependent on ADC, small changes in ADC can make a significant change in DWI contrast. This result the significant increase of CR in sDWI images as compared to DWI.

The present study demonstrates that, although the higher *b*-value sDWI increases the contrast between lesion and normal tissue, the sADC shows similar contrast for *b*-1000, *b*-1500 and *b*-2000. This could be the due to small cohort size of the patient with different Gleason score, consistent with results in other studies (24,25). ADC computed from high b-value DWI has been shown to be more accurate in distinguishing prostate lesions from benign and normal tissues (26,27).

Our initial investigation suggests that the ADC values corresponding to higher *b*-value DWI can be computed using a log-linear relationship derived from lower *b*-values (*b*≤1000). Moreover, this computational method can also be manipulated to determine optimized *b*-values to create ADC maps. The synthetic ADC technique could be a useful tool to provide optimized image contrast for quantitative DW MR imaging applications in oncology where ADC is routinely used in clinical practice.

## Data Availability

This is a retrospective study. The data that support the findings of this study are available on request from the corresponding author PS. The data are not publicly available.

## Notes

### Competing Interest Statement

The authors have declared no competing interest.

### Funding Statement

no external funding was received

### Author Declarations

All relevant ethical guidelines have been followed and any necessary IRB and/or ethics committee approvals have been obtained.

Any clinical trials involved have been registered with an ICMJE-approved registry such as ClinicalTrials.gov and the trial ID is included in the manuscript.

